# Photon-counting computed tomography for phantom-less quantitative measures of musculoskeletal tissues

**DOI:** 10.64898/2026.07.22.26358681

**Authors:** Steven K. Boyd, Niklas A. Lackner, Anna-Maria Liphardt, Matthias S. May, Georg Schett, Michael Uder, Klaus Engelke

**Affiliations:** McCaig Institute for Bone and Joint Health, University of Calgary, 3280 Hospital Drive NW, T2N 4Z6, AB, Canada; Department of Biomedical Engineering, Schulich School of Engineering, University of Calgary, AB, Canada; Department of Radiology, Cumming School of Medicine, University of Calgary, AB, Canada; Institute of Radiology, Friedrich-Alexander-Universität Erlangen-Nürnberg and Uniklinikum Erlangen, Erlangen, Germany; Imaging Science Institute, Uniklinikum Erlangen, Erlangen, Germany; Department of Medicine 3 - Rheumatology and Immunology, Friedrich-Alexander-Universität Erlangen-Nürnberg and Uniklinikum Erlangen, Erlangen, Germany; Deutsches Zentrum für Immuntherapie (DZI), Friedrich-Alexander-Universität Erlangen-Nürnberg and Uniklinikum Erlangen, Erlangen, Germany

**Keywords:** computed tomography, bone, adipose, muscle, bone mineral density, photon-counting CT, dual-energy CT, material decomposition, virtual monoenergetic image

## Abstract

The advent of photon-counting computed tomography (PCCT) provides new opportunities to quantitatively measure musculoskeletal tissues such as bone, muscle and adipose because of the intrinsic use of spectral imaging. We aimed to evaluate the accuracy of measuring these tissues by PCCT under a range of scan protocols and compared our results to the current standard dual-energy CT (DECT). Phantoms containing inserts ranging from 50 to 200 *mg/cm*^3^ of calcium hydroxyapatite (HA) for estimating bone mineral density (BMD), and another phantom containing inserts for muscle and adipose tissues were scanned on PCCT and DECT at 120 and 140 kVp. We created virtual monoenergetic images (VMI) at energy levels from 40 keV to 190 keV for quantitative analyses. The averaged linear attenuation of phantom inserts was compared to theoretical values calculated from standardized attenuation profiles. Material decomposition using VMIs was compared to known HA concentration inserts to determine optimal image pairs for BMD measurement, notably without the need of phantom calibration. For most VMI energy levels the attenuation error was <1% for BMD at both 120 kVp and 140 kVp by PCCT compared to errors of <2% by DECT. The linear attenuation errors were <2.5% for muscle and <3.0% for adipose and results were similar for PCCT and DECT. Generally, errors were highest for low energy VMIs. Material decomposition using VMI pairs with a low energy at 50 or 60 keV and high energy between 150 and 190 keV produced calibration phantom-free estimates of BMD with <1% error. Results were similar for PCCT and DECT at 120 and 140 kVp. PCCT provides an accurate estimate of bone, muscle and adipose attenuation, and using material decomposition, estimations of BMD can be obtained without the need of phantom calibration. Given that multi-energy imaging is intrinsic for PCCT this provides new possibilities for opportunistic screening in patients who get PCCT scans for other clinical indications.

## Introduction

Computed tomography (CT) is a highly utilized diagnostic tool with millions of scans performed annually worldwide. Most CT scans are used for qualitative clinical evaluation, with only a small proportion incorporating quantitative data even though CT is well-suited for that purpose. This is an untapped resource, particularly in the musculoskeletal field. For example, quantitative measures of bone mineral density (BMD) could support diagnoses of osteoporosis, or measures of muscle quality and related intramuscular fat could detect sarcopenia. Quantitative measures from CT imaging would support opportunistic individual assessments as well as implementation of pragmatic clinical trials for real-world testing of medical treatments and interventions.

Current opportunistic analyses frequently do not calibrate for quantitative measures and instead report in CT numbers (Hounsfield Units; HU), which are well known to be affected by machine calibration and acquisition settings (e.g., X-ray tube energy, table positioning) (8). Even if calibration is performed, conventional single-energy CT imaging cannot dissociate bone marrow fat from trabecular bone, resulting in a bias in quantitative BMD measurements due to the attenuation properties of fat that are lower than those of the reference material water. This results in negative HU for fat that biases the total used for a measure of BMD. For example, in the lumbar spine the percent of marrow fat is substantial, up to 20% (10; 13; 29), and typically increases with age resulting in an apparent BMD decrease (14; 18). This results in large errors that are approximately 10–30% at 80 kV and 20–40% at 120 kV (14). In elderly subjects bone marrow of the hip and distal forearm is almost 100% fat resulting in even larger BMD accuracy errors.

Multi-energy imaging can be used to distinguish mixtures of tissues (i.e., bone and marrow fat) by the method of material decomposition so that quantitative analyses can be performed (20; 15; 3). For nearly two decades, dual-energy CT (DECT) has been routinely available in the clinic and can provide fat-corrected images of bone, but has limited applicability for opportunistic analyses because it is not frequently indicated clinically. The recent introduction of photon-counting CT (PCCT) (12; 11; 26; 22) offers exciting new possibilities for clinical imaging (21; 4; 12; 28). Compared to standard DECT it requires a relatively lower dose and provides increased spatial and spectral resolution. Importantly, in contrast to the energy integrating detector used in most DECT scanners, the spectral information is *intrinsically* collected by the novel photon-counting detector, which means that all clinical exams, even when multi-energy imaging is not necessary, can potentially be used for quantitative imaging. Early exploration of PCCT for musculoskeletal research has focused on its spatial resolution capabilities, which have shown promising results for detailing bone structure at the radius (5; 25; 24; 23; 30). It is not yet clear to what degree these benefits of resolution extend to axial skeletal sites that are key for osteoporosis assessments (i.e., spine and hip). On the other hand, the potential for PCCT to generate phantom-less, fat-corrected images of bone by PCCT provides new opportunities for opportunistic analysis. Virtual monoenergetic images (VMIs) can be generated using a linear combination of the basis material images (e.g., iodine and water) that are derived from the low (L) and high (H) spectra in the projection domain. The VMIs represent simulated attenuation at a given photon energy, akin to a monoenergetic acquisition, that enables post-processing for material decomposition in the image domain (31).

The objective of this study is two-fold: First, to compare the accuracy of PCCT for measuring the CT values [HU] of bone, muscle and adipose from VMIs against theoretical values calculated from the established attenuation profiles provided by the National Institute of Standards and Technology (NIST). Second, to characterize which VMI energy selection pairs are best suited for material decomposition to perform phantom-less estimation of bone mineral density. All our PCCT quantitative measures will be compared to the well-established DECT method.

## Methods & Materials

### Phantoms

Three CT phantoms (QRM GmbH, Moehrendorf, Germany) were used for image acquisition and analysis (Fig. 1):

1. European Spine Phantom (ESP; s/n ESP-143)
2. Bone Density Calibration (BDC; s/n HQ-01-02)
3. Electron Density Phantom (EDP; s/n EDP-04-09)

**Figure 1.**
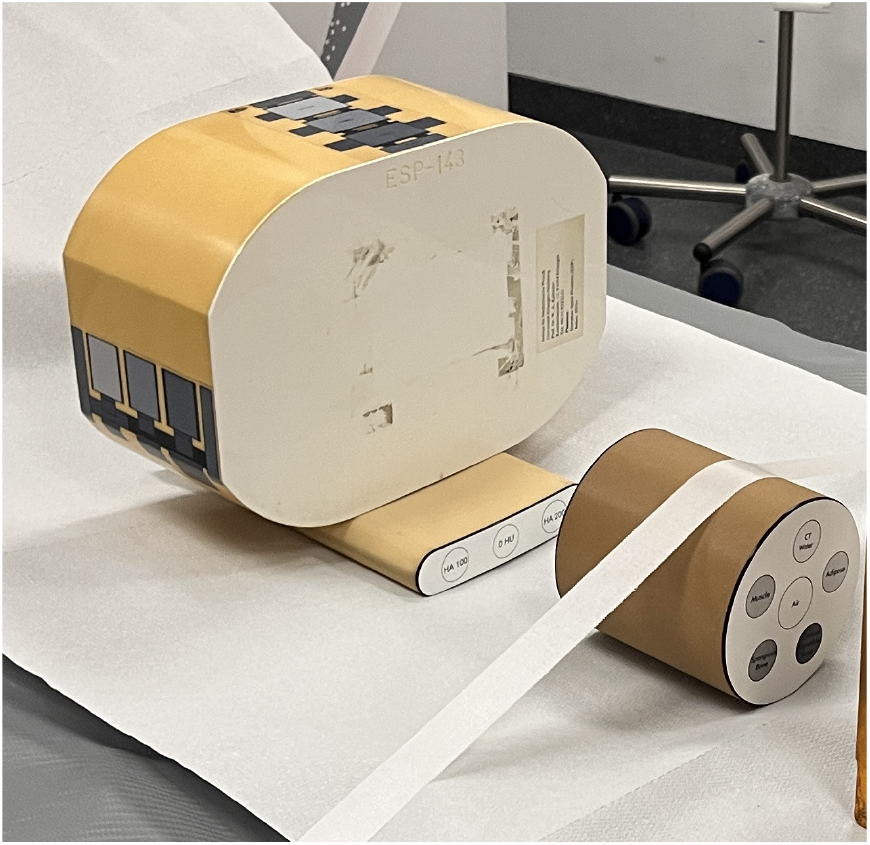
The European Spine Phantom (ESP) with a Hip QC Phantom (BDC) placed underneath and the Electron Density Phantom (EDP) on the computed tomography scanner bed.

The ESP and the BDC phantoms have inserts with defined concentrations of calcium hydroxyapatite (HA; *Ca*_10_(*PO*_4_)_6_(*OH*)_2_) (Table 1) and the EDP has inserts for materials of adipose, muscle, CTWater^©^, cortical bone and spongiosa bone that are defined by the International Commission on Radiation Units and Measurements (ICRU) (1; 2). The manufactured accuracy of these certified values is <1% (17). The composition of ICRU-defined materials in the phantoms (Table 2) were used to derive theoretical mass attenuation curves representing linear attenuation from 40 to 200 keV (the relevant clinical range), which served as the reference for comparison with measured values. The NIST^1^ data are provided for some materials or can be generated using the NIST XCOM^2^ software.

**Table 1.**
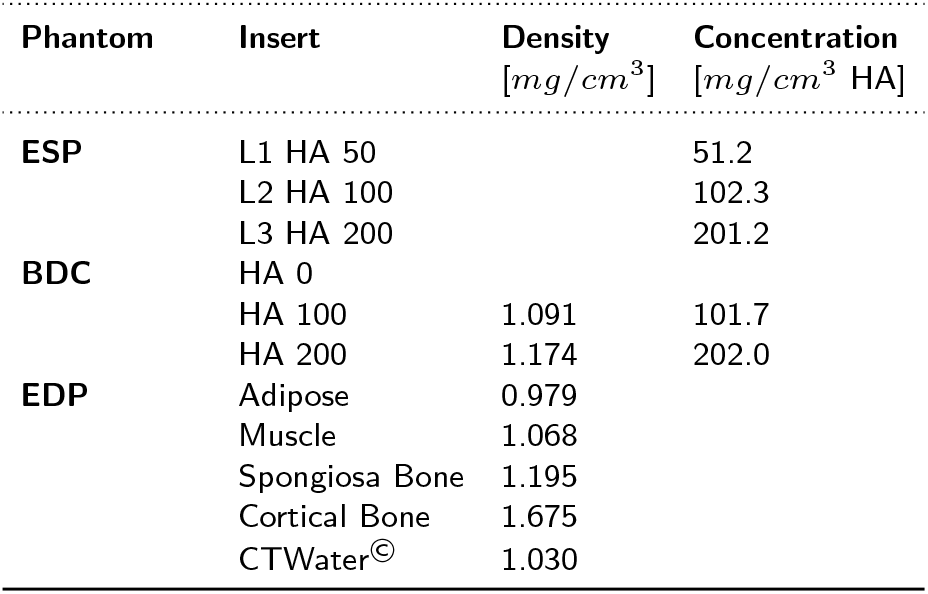
The nominal physical densities and concentrations of calcium hydroxyapatite (HA) as certified by QRM for the European Spine Phantom (ESP), Bone Density Calibration (BDC) phantom, and Electron Density Phantom (EDP). For the EDP, certified physical densities are also listed for adipose, muscle, spongiosa bone, cortical bone and CTWater.

| Phantom | Insert | Density<br>[mg/cm<sup>3</sup>] | Concentration<br>[mg/cm<sup>3</sup> HA] |
| --- | --- | --- | --- |
| ESP | L1 HA 50 |  | 51.2 |
|  | L2 HA 100 |  | 102.3 |
|  | L3 HA 200 |  | 201.2 |
| BDC | HA 0 |  |  |
|  | HA 100 | 1.091 | 101.7 |
|  | HA 200 | 1.174 | 202.0 |
| EDP | Adipose | 0.979 |  |
|  | Muscle | 1.068 |  |
|  | Spongiosa Bone | 1.195 |  |
|  | Cortical Bone | 1.675 |  |
|  | CTWater<sup>®</sup> | 1.030 |  |

**Table 2.** The fractional elemental content (by mass) of key materials used for calculating mass attenuation properties as defined in ICRU reports (1; 2) for adults. Hydroxyapatite is defined as *Ca*_10_(*PO*_4_)_6_(*OH*)_2_.

| Material | H | C | N | O | Ca | Na | S | Cl | Mg | P | K | Fe | $\rho$ |
| --- | --- | --- | --- | --- | --- | --- | --- | --- | --- | --- | --- | --- | --- |
| Units | % | % | % | % | % | % | % | % | % | % | % | % | $g/cm^3$ |
| Water | 11.2 |  |  | 88.8 |  |  |  |  |  |  |  |  | 1.000 |
| CTWater<sup>®</sup> | 8.1 | 69.9 | 2.0 | 18.0 | 1.9 |  |  | 0.1 |  |  |  |  | 1.030 |
| Spongiosa | 8.5 | 40.4 | 2.8 | 36.7 | 7.4 | 0.1 | 0.2 | 0.2 | 0.1 | 3.4 | 0.1 | 0.1 | 1.180 |
| Cortical | 3.4 | 15.5 | 4.2 | 43.5 | 22.5 | 0.1 | 0.3 |  | 0.2 | 10.3 |  |  | 1.920 |
| Hydroxyapatite | 0.2 |  |  | 41.4 | 39.9 |  |  |  |  | 18.5 |  |  | 3.225 |
| Muscle | 10.2 | 14.3 | 3.4 | 71.0 |  | 0.1 | 0.3 | 0.1 |  | 0.2 | 0.4 |  | 1.050 |
| Soft tissue |  |  |  |  |  |  |  |  |  |  |  |  |  |
| male | 10.5 | 25.6 | 2.7 | 60.2 |  | 0.1 | 0.3 | 0.2 |  | 0.2 | 0.2 |  | 1.030 |
| female | 10.6 | 31.5 | 2.4 | 54.7 |  | 0.1 | 0.2 | 0.1 |  | 0.2 | 0.2 |  | 1.020 |
| Adipose |  |  |  |  |  |  |  |  |  |  |  |  |  |
| adult #1 | 11.2 | 51.7 | 1.3 | 35.5 |  | 0.1 | 0.1 | 0.1 |  |  |  |  | 0.970 |
| adult #2 | 11.4 | 59.8 | 0.7 | 27.8 |  | 0.1 | 0.1 | 0.1 |  |  |  |  | 0.950 |
| adult #3 | 11.6 | 68.1 | 0.2 | 19.8 |  | 0.1 | 0.1 | 0.1 |  |  |  |  | 0.930 |

### Data Acquisition

Scans were performed at the Department of Radiology of the Erlangen University Hospital on a dual-source PCCT (NAEOTOM Alpha, VB20A, Siemens Healthineers AG, Forchheim, Germany) and a twin beam DECT (SOMATOM X.cite, VB20A, Siemens Healthineers AG, Forchheim, Germany).

The PCCT uses a photon counting detector (PCD) with multiple energy thresholds from which spectrally separated image datasets are reconstructed. Four energy thresholds are mixed and decorrelated to reduce the correlations in noise from the images (9) producing a low (L) and high (H) image dataset. PCCT scans were performed with tube voltages of 120 kV and 140 kV. Exploratory scans were also acquired at 90 kV; however, the resulting spectral separation was insufficient for stable material decomposition and these data were therefore not included in the subsequent analyses. PCCT acquisition parameters used a spiral pitch of 0.8, slice width of 0.4 mm, and reconstruction into a 512 by 512 matrix (nominally 0.28 mm in-plane voxel size) using filtered back-projection (Br40f*\*3).

The DECT scans were performed in TwinBeam Dual Energy mode where a single X-ray source has a split beam with gold (Au) and tin (Sn) filters placed in the beam path resulting in two distinct spectra. Scans were performed at 120 kV (AuSn120kV) and 140 kV (AuSn140kV). Acquisition parameters were set to a spiral pitch of 0.45, slice width of 0.6 mm, and reconstruction into a 512 by 512 matrix (nominally 0.22 to 0.40 mm in-plane voxel size) using filtered back-projection (Qr40f).

Both PCCT and DECT scans had tube current and exposure adjusted to ensure a consistent CTDI of 11.8 mGy. Reconstructions of VMIs were created from 40 keV to 190 keV in steps of 10 keV (syngo.via, Siemens Healthineers AG, Forchheim). VMIs were generated by the vendor’s spectral reconstruction software from the spectrally separated datasets; VMI synthesis can be expressed either via basis-material images or as a weighted combination of low/high images (20). On the PCCT system an additional reconstruction (T3D) that most closely emulates a conventional CT with an energy integrating detector was generated.

All data had volumes of interest (VOI) defined that were approximately 60% of the diameter of each cylindrical insert and covered at least 80 slices. These VOIs were used to calculate the mean CT number for each insert as a function of tube spectrum (kV) and VMI energy (keV).

### Estimation of linear attenuation

For a VMI at a given energy (keV) we estimated the linear attenuation, 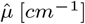, of a mixture of materials *m*_*N*_ having a partial volume fraction *f*_*m*_ using the following equation:

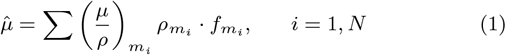

where (*µ/ρ*) [*cm*^2^ *· g*^*−*1^] is the mass attenuation, *ρ* [*g/cm*^3^] is the ICRU-defined mass density of material, *m*_*i*_, and the volume fraction 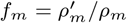 [%] is subject to the volume constraint:

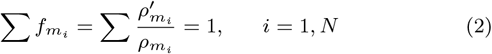

The mass concentration of material *m* is denoted as *ρ*^*′*^. The mass attenuation was based on the NIST values for ICRU materials. For compounds it was estimated using the NIST XCOM online calculator.

It is useful to use an example for illustrative purposes. If we have an HA insert in a phantom, which is a mixture of HA and water-equivalent material (CTWater^©^), *w*, then we expand Equation 1 to consider the mixture of the HA and water equivalent material as follows:

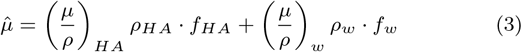

Taking the BDC phantom 200 HA insert (Table 1) as an example, which has a nominal HA concentration of *ρ*^*′*^ = 202.0 *mg HA/cm*^3^, we calculate the volume fraction of HA as *f*_*HA*_ = 202.0*/*3225 = 0.0626 and use Equation 2 solve for the volume fraction of water as *f*_*w*_ = 1 *− f*_*HA*_ = 0.9374. At 100 keV the mass attenuation for HA and *w* are interpolated (log-log) from the mass attenuation curves:

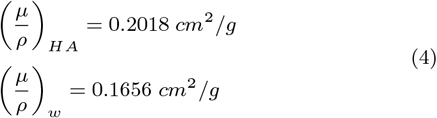

and so we calculate the estimated linear attenuation as per Equation 3 and find 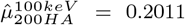. We convert this linear attenuation coefficient to a CT number [HU] using the standard equation:

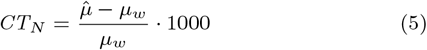

where the mass attenuation of water, (*µ/ρ*)_*w*_, at 100 keV is 0.1707 *cm*^2^*/g*. Since the ICRU mass density of water *ρ*_*w*_ is 1.0 *g/cm*^3^ and the volume fraction *f*_*w*_ = 1.0 then the linear attenuation of pure water, *µ*_*w*_, by Equation 1 is 0.1707*/cm*. The resulting calculated *CT*_*N*_ is 178 HU compared to the mean value we sampled from the VOI was also 178 (140 kV tube spectrum), resulting in a difference of 0 HU and an error of linear attenuation *µ* of 0.0%.

### Image-Based Material Decomposition

We performed image-based *2-material* decomposition with a pair of VMIs representing a low (*L*) and high (*H*) attenuation image. The *2-material* decomposition was performed using Equation 1, which can be expanded:

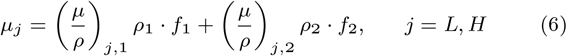

Since *f*_1_ *· ρ*_1_ = *ρ*^*′*^_1_ and *f*_2_ *· ρ*_2_ = *ρ*^*′*^_2_ we can write:

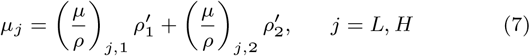

We express the solution of Equation 7 in matrix form (*Ax* = *b*) and solve it for all voxel pairs in the *µ*_*L*_ and *µ*_*H*_ images using linear algebra:

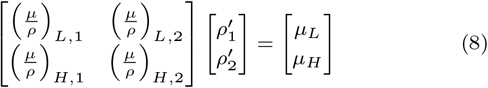

The voxel-wise solution for mass concentrations of materials used in the decomposition are stored in an image map of *ρ*^*′*^_1_ and *ρ*^*′*^_2_ . In order to solve Equation 8 the low and high CT images must be converted from Hounsfield Units to linear attenuation, *µ*, by Equation 5.

Theoretically any two materials can be chosen provided sufficient spectral separation exists, but here we are interested in HA and water because our rods are composite mixtures of these materials. All combinations of VMIs from 40 keV to 190 keV were tested for *2-material* decomposition from either PCCT or DECT images. Errors were represented as a percent error of *µ*:

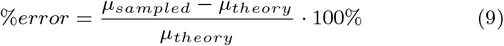

Software for estimating linear attenuation (ogoCalculateLinearAttenuation) and material decomposition (ogoMaterialDecomposition) is available from the Bone Imaging Laboratory repository (GitHub Ogo).

## Results

Using the ESP phantom measured at the three levels (L1, L2, and L3) we sampled the mean CT number from a subset of VMIs at 50, 60, 70, and 80 keV, as well as the T3D image (Figure 2) generated from tube voltages at 90 kV, 120 kV, and 140 kV acquisitions. These data illustrate that for the VMI images there is a strong dependence on the keV, but relatively little dependence on the X-ray tube voltage. In contrast, the T3D image, which most closely represents a conventional CT image, shows a strong dependency on X-ray tube voltage. This result follows closely to data previously shown by McCollough and colleagues (22) and is at the heart of why using CT values as a surrogate for bone density in conventional CT is problematic.

**Figure 2.**
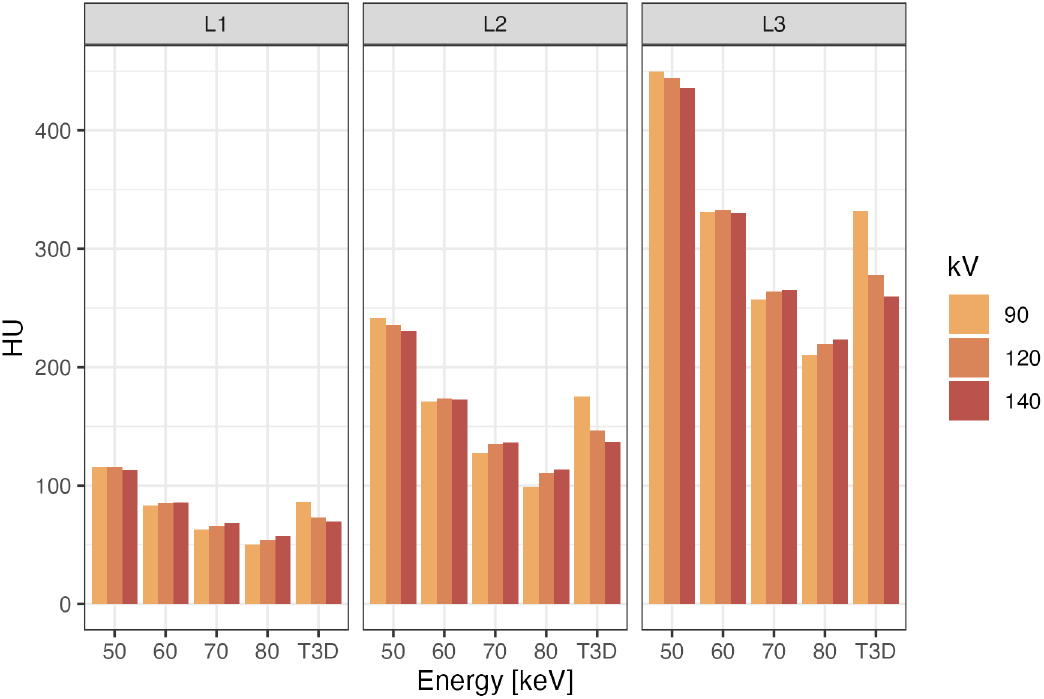
CT number dependence on tube voltage for calcium hydroxyapatite (ESP; L1 = 51.2, L2 = 102.3, L3 = 201.2 *mgHA/cm*^3^) for virtual monoenergetic images (VMIs) and the polychromatic T3D images collected on the PCCT at 90, 120 and 140 kV.

The percent accuracy error of estimated linear attenuation was determined from the VMI images in the range 40 keV to 190 keV for PCCT and DECT measured at 0, 100, and 200 *mg/cm*^3^ concentrations of HA (Figure 3) and ICRU-defined adipose, muscle and spongiosa bone (Figure 4). The absolute CT number used to estimate the linear attenuation for PCCT and DECT is provided for the ICRU-defined materials and three HA concentrations representing the physiological range of trabecular bone (Table 3).

**Table 3.**
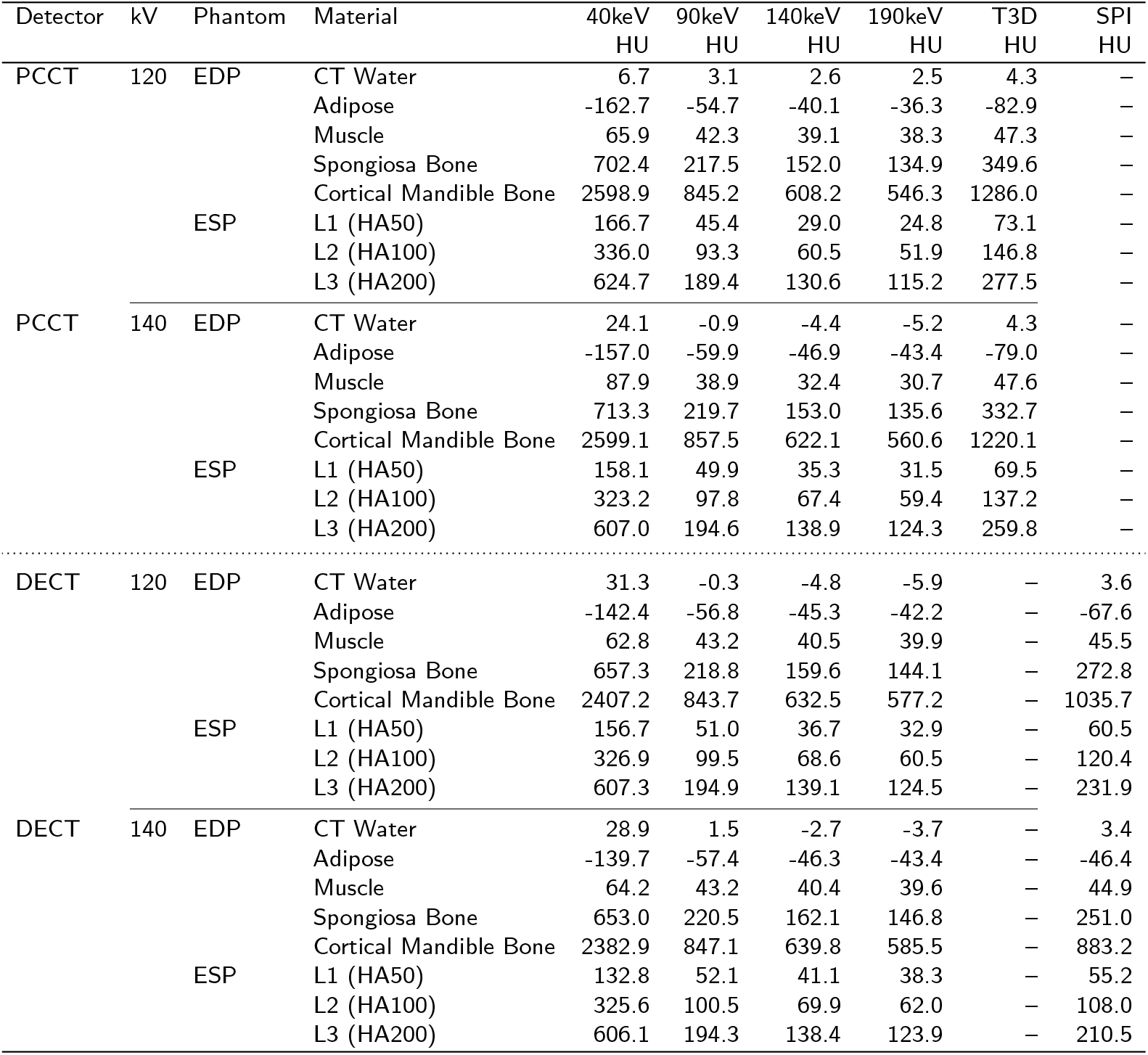
CT number measured by photon-counting CT (PCCT) and dual-energy CT (DECT) at 120 and 140 kV for a subset of VMIs between 40 keV and 190 keV in addition to the Threshold + 3D image (T3D) from the PCCT and spiral scan mode image (SPI) from DECT. The mean Hounsfield Unit (HU) is shown for all the inserts in the Electron Density Phantom (EDP) and the European Spine Phantom (ESP).

**Figure 3.**
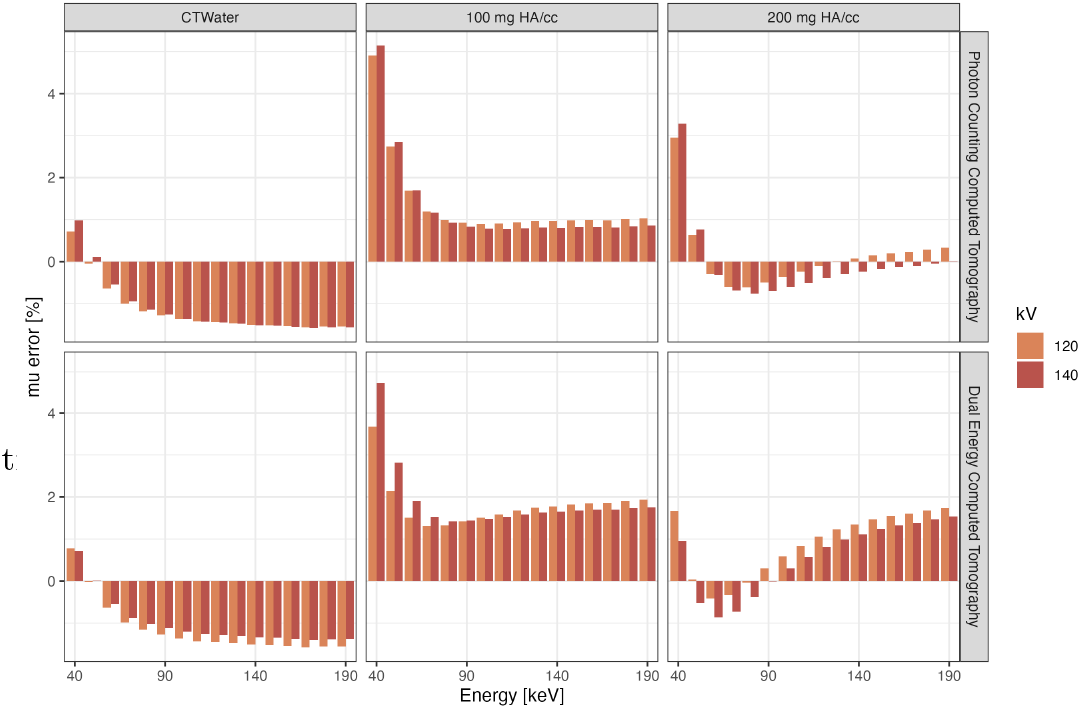
Photon-counting CT and dual-energy CT estimates percentage errors in linear attenuation for virtual monoenergetic images (VMIs) ranging from 40 keV to 190 keV. The estimates are based on the BDC phantom for CT-equivalent water (CTWater^©^) and calcium hydroxyapatite inserts at 100 *mgHA/cm*^3^ and 200 *mgHA/cm*^3^.

**Figure 4.**
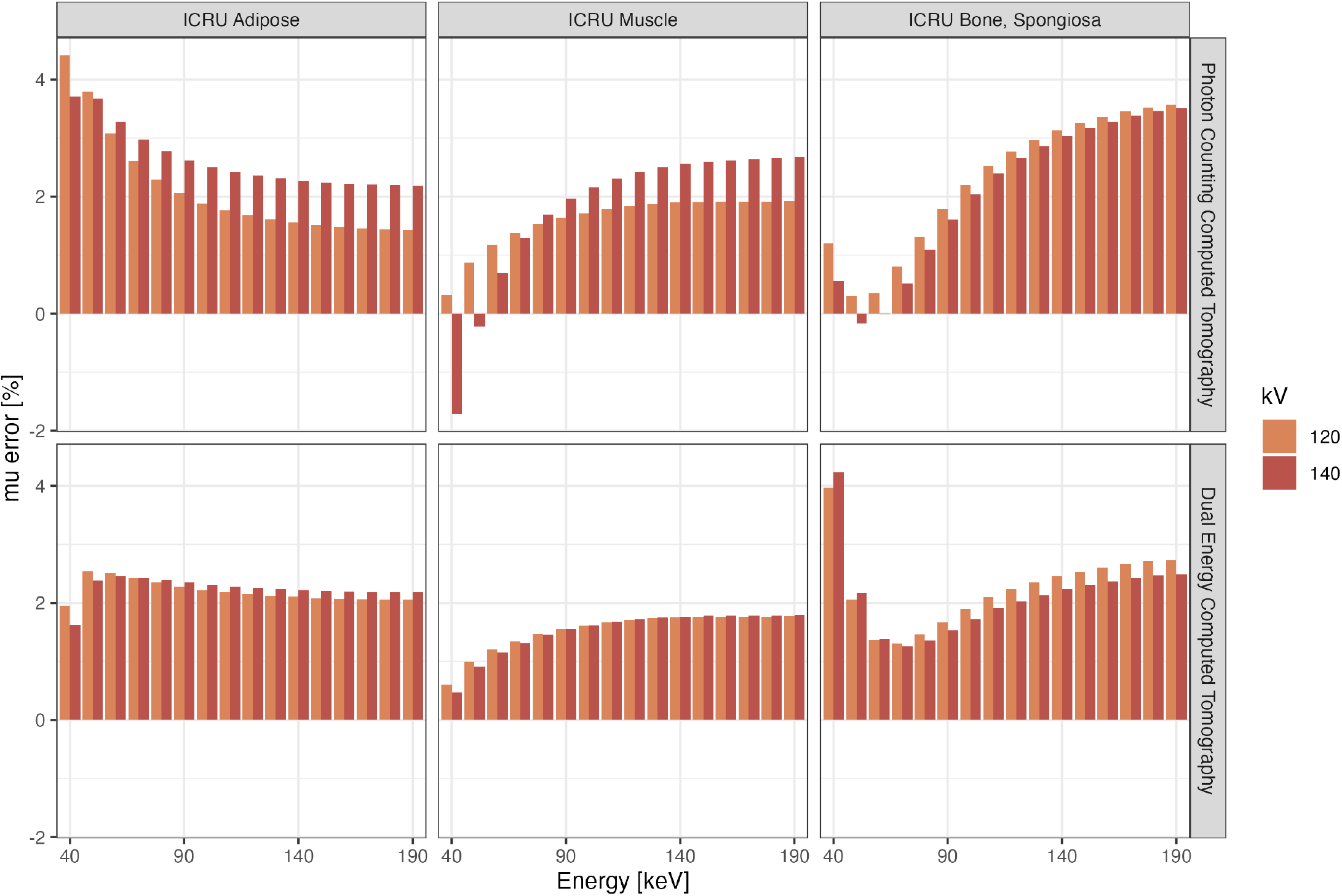
Photon-counting CT and dual-energy CT percentage errors in linear attenuation for adipose, muscle, and spongiosa bone for virtual monoenergetic images (VMIs) ranging from 40 keV to 190 keV. The estimates are based on the electron density (EDP) phantom with ICRU-defined adipose, muscle and spongiosa bone.

We performed material decomposition using HA and CTWater^©^ as reference materials for all available combinations of VMIs from 40 keV to 190 keV, which are the two materials comprising HA rods in the phantom. Our output is a HA density map from which we calculated the percent error of the measured HA concentration, *ρ*_*HA*_, relative to the calibration phantom certified values. The errors were determined for both PCCT and DECT (Figure 5) and ranged depending on the selection of low and high image, with excellent accuracy typically when the low (L) VMI was 50 to 60 keV and the high (H) VMI was 160 keV or greater.

**Figure 5.**
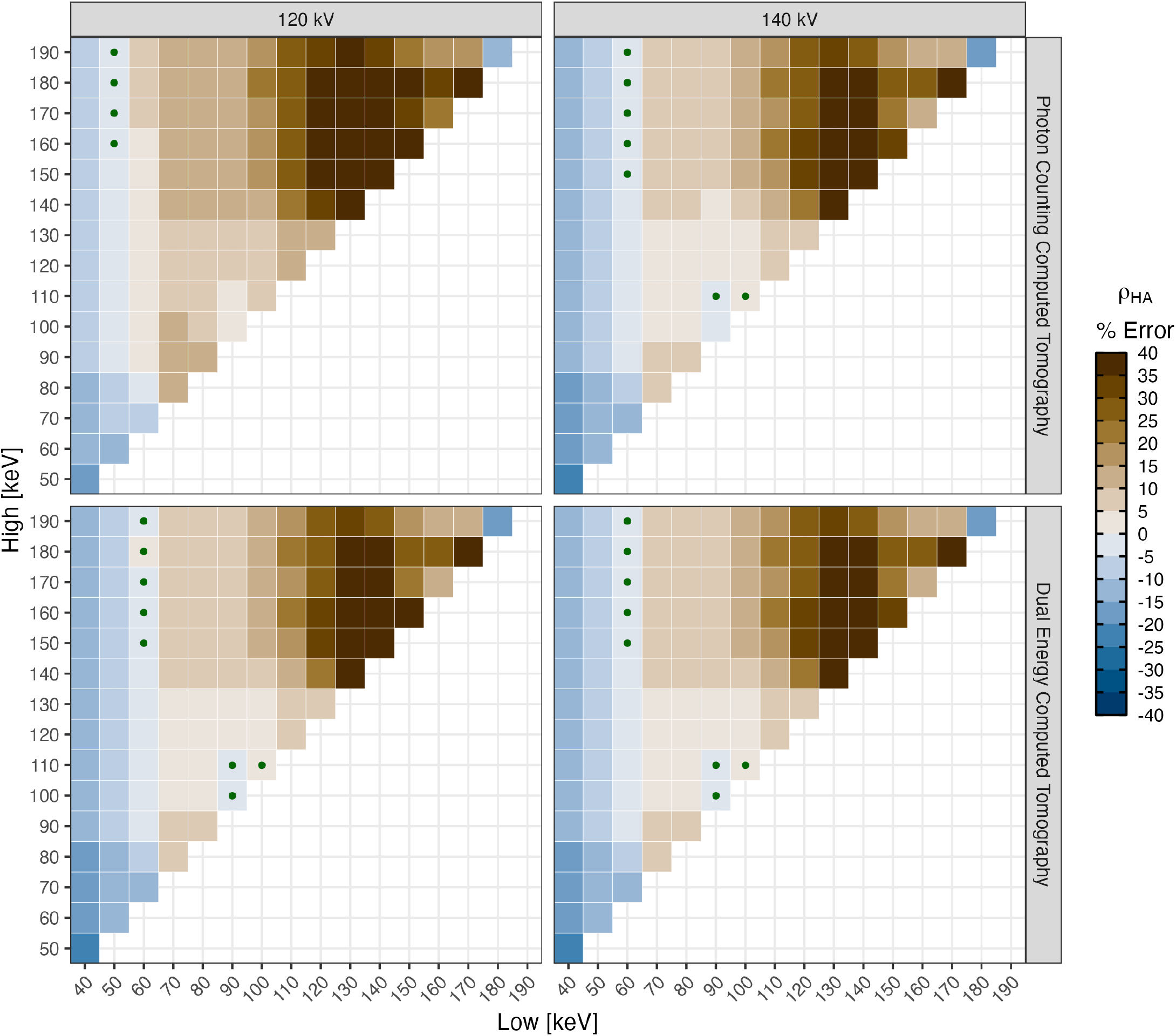
Photon-counting CT and dual-energy CT estimates of the ESP L3 using *2-material* decomposition from pairs of VMIs generated from tube potentials of 120 kV and 140 kV. Contours are expressed as a percent error of the estimated concentration calcium hydroxyapatite (HA) compared the certified phantom values. VMIs pairs resulting in less than 1% error are indicated with a green circle. These measures were phantom-less estimates of HA.

## Discussion

This study has demonstrated the excellent accuracy of PCCT for quantitative assessment of musculoskeletal tissues relevant to bone health. In particular, BMD can be quantified without external calibration, which is highly relevant for opportunistic screening. We found excellent linear attenuation estimates to predict HA and soft tissue concentrations by PCCT based on certified CT phantoms. For VMIs greater than 60 keV linear attenuation was consistently within 2% of theoretical values, with moderately lower errors by PCCT compared to DECT. Subsequently, we showed that material decomposition to provide quantitative maps of BMD produced PCCT prediction errors less than 1% when appropriate low and high VMIs pairs were chosen. This is an important finding because using VMIs in post-processing combined with physics-based priors (i.e., NIST attenuation) enables accurate phantom-less estimates of BMD maps.

The sampled linear attenuation from PCCT and DECT images differed from the theoretical values as a function of the VMI energy, with the greatest errors at the lower energies. This trend is expected because these VMIs are furthest from the effective energy of the acquisitions (Table 4) and because at the low energies photon loss leads to increased noise (20). Furthermore, the use of iodine and water as the base pair by the manufacturer software for generating VMIs may have contributed to the error. Consequently, since the VMIs are linear combinations of the low and high spectra, the difference in their effective energies influence the resulting VMI attenuation maps. The effective energy of the low spectra for PCCT at 120 kV and 140 kV is approximately 60 keV, and for DECT it is in the mid 70’s keV range. The errors in linear attenuation for both PCCT and DECT are smaller for higher VMI energies. The differences in effective energy of DECT and PCCT is due to their different detector response (energy weighting), which gives the PCCT images a lower effective energy compared to DECT for the same incident X-ray spectrum. It is likely this difference is the reason why there is a consistently lower attenuation prediction error for PCCT compared to DECT.

**Table 4.**
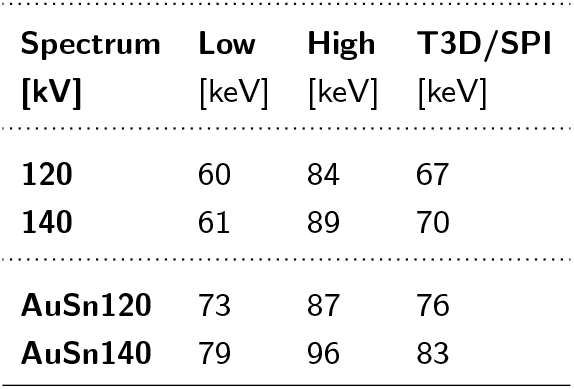
The effective energy of the low (L), high (H) and total (T3D for PCCT or SPI for DECT) acquisitions as a function of tube spectrum.

Material decomposition for estimating BMD in the European Spine Phantom was most accurate if appropriate VMIs image pairs were chosen. The VMI pairs that worked best were those with sufficient spectral separation so there is distinct information in each image, which is why we find errors are greatest if both are low or both are high. The selection of the low energy VMI had the greatest impact on error sensitivity. We found that 60 keV consistently provided good results whereas the high energy VMI could be any within the range of 150 keV to 190 keV to provide the lowest errors. Selecting the lowest energy VMI (i.e., 40 keV) resulted in increased noise, so there is a trade-off where the benefits of spectral separation are balanced against the increased noise at lower VMIs, with 50 keV being a compromise. Notably, both the PCCT and DECT produced similar results, particularly for the 140 kV X-ray tube energy acquisitions where the best combinations were nearly identical. This result is important because it suggests both spectral approaches have the ability to incorporate post-processing (i.e., material decomposition from vendor-based VMIs) to estimate BMD measurements purely using physics-based priors rather than the traditional approach of depending on phantom-based image calibration.

The vendor-derived VMIs were generated for iodine and water basis materials, which are not optimal for our targeted assessment of musculoskeletal tissues, particularly BMD. The VMIs are most quantitatively accurate to assess materials that are close to the chosen basis materials (e.g., iodine-water, calcium-water). The default acquisition mode of the PCCT we used is an iodine-water base pair, but our future work is exploring whether a calcium-water base pair can provide more accurate measures. If so, it is possible that the excellent measurement accuracy we found here for quantitative tissue densities may be even further be improved. If possible in the future, selecting the base material pair closest to the tissues of interest is recommended.

Others exploring the use of PCCT for musculoskeletal applications have focused on its improved resolution for the assessment of bone microarchitecture compared to conventional CT. Using HR-pQCT as a gold-standard, it has been demonstrated that PCCT-derived microarchitectural parameters are highly correlated (5; 24).

However, the focus to date has been on the distal radius primarily because the chosen gold-standard HR-pQCT can only perform measurements at peripheral sites. The axial skeleton (i.e., hip, spine) is of particular importance because of its relevance to osteoporotic fracture, but it is not yet known how accurately microarchitecture can be measured, which is likely more challenging due to effects of beam hardening. Unfortunately, spectral information when acquiring images at the highest resolution is currently not available, yet incorporating that into microarchitectural assessments in the future could have the benefit of ensuring improved segmentation protocols (i.e., thresholding). Without spectral information, the outcome measure in HU is susceptible to acquisition settings such as X-ray tube voltage, even if performed at high resolution. Further, there is a confounding influence of marrow fat, which is prominent in the vertebral body. Ideally, integrating high resolution imaging with material decomposition to provide accurate quantitative images (i.e., density) will hopefully be possible in the future, which would increase the potential benefits of PCCT for bone microarchitecture.

One of the challenges of material decomposition using VMIs in clinical practice is that they may not always be available for post-processing. In contrast to DECT where spectral information is only collected if clinically indicated, PCCT collects spectral information intrinsically and independent of clinical indication. However, if vendor-based VMIs are not generated as part of the clinical protocol, that information will be lost and unavailable for post processing.

That would be a lost opportunity, especially since there is no additional radiation to the patient or added exam time. The only cost of retaining VMIs is the nominal computational expense and PACS storage requirements. It is recommended that PCCT protocols be standardized to include at least two VMI reconstructions (i.e., 60 and 160 keV) from each acquisition so that post-processing analysis is possible. This would enable all PCCT scans to be used opportunistically and maximize the clinical benefits of these radiological exams.

There are limitations to our study that should be noted. First, the calibration phantoms we used contain homogeneous mixtures of known concentrations of HA and ICRU-defined materials, but phantom designs have continued to improve since the CT phantoms we used were constructed. It is possible that a small error is introduced due to the difference between the composition of the phantom inserts and the physics-based mass attenuation curves obtained from NIST. Second, we tested material decomposition with inserts of HA and water mixtures, whereas a mixture of HA, water and adipose would be more relevant for the skeleton. Similar to conventional single-energy QCT, the accuracy of BMD results obtained with the method introduced in this study will be impacted by the presence of marrow fat. We are not aware of phantoms that contain mixtures of HA and adipose, but are planning to construct and test these in the future. Third, our material decomposition was for two materials, but there are cases where it may be important to employ decomposition into three materials (16; 27; 7) and for a wide variety of materials (19). For example, bone marrow edema in an anterior cruciate ligament injured knee may be important to distinguish from surrounding trabecular bone and marrow fat. We are not aware of phantoms for testing decomposition of three materials, which is why previous implementation of three-material decomposition has used qualitative comparisons to other imaging modalities (6). In general, more sophisticated CT phantoms will be important to explore in the future, coupled with validation against real human tissue. Fourth, our selection of optimal VMI pairs are dose-dependent and specific to the HA concentrations we tested, so different optimal pairs may need to be determined for specific testing conditions. Also, they were determined for a single vendor (Siemens Healthineers) and may not generalize to other CT systems. Fifth, accurate prediction of attenuation properties from mass attenuation curves is important to our physics-based approach and so in the clinical range of 30 keV to 200 keV where these curves are exponential we found that *log-log* interpolation with smoothing worked better than *linear* or *cubic* interpolation options. The interpolation is important because it can have a direct effect on the phantom-less BMD estimates from material decomposition. Lastly, accurate BMD measures require that the CT scanner is calibrated so that the CT value of water is zero (for any tube voltage setting). In both scanners a small offset was observed (see Table 3). Typically in conventional single-energy QCT such an offset is subtracted from the CT values of bone during the BMD calibration, which may have further improved BMD accuracy. However, it is not entirely clear whether this procedure can also be applied to the VMIs and their combination to obtain BMD values, thus no water offset correction was performed in this study.

## Conclusions

PCCT imaging allows characterization of musculoskeletal tissue attenuation, and material decomposition can be used to estimate the concentration of HA in calibration inserts with errors less than 2%, which enables *phantom-less* BMD calibration in the clinical setting. In this experimental setting, PCCT produced modestly lower errors than DECT. A major advantage of PCCT is that spectral information is intrinsically available in routine acquisitions, thus it is recommended that VMI pairs be archived in all exams to enable *post hoc* quantitative analysis and derive maximum benefit from this new technology.

## Data Availability

Data is available upon reasonable request.

## Acknowledgments

The authors would like to thank the support of Thomas Flohr and Sebastian Faby for their input and advice.

## Authors’ Contributions

SB, NL, AML, MM, KE (a) conceptualized the study (b) performed image analysis (c) conducted the data analysis (d) drafted the manuscript. All authors provided input and approved the final version.

## Data Availability

Data is available upon reasonable request. Code is available in the public Ogo GitHub repository:

https://github.com/Bonelab/Ogo

## Footnotes

1 https://physics.nist.gov/PhysRefData/XrayMassCoef/tab4.html

2 https://www.nist.gov/pml/xcom-photon-cross-sections-database

